# Impact of BNT162b first vaccination on the immune transcriptome of elderly patients infected with the B.1.351 SARS-CoV-2 variant

**DOI:** 10.1101/2021.05.11.21256862

**Authors:** Ludwig Knabl, Hye Kyung Lee, Manuel Wieser, Anna Mur, August Zabernigg, Ludwig Knabl, Simon Rauch, Matthias Bock, Jana Schumacher, Norbert Kaiser, Priscilla A. Furth, Lothar Hennighausen

**Affiliations:** TyrolPath, Zams, Austria; National Institute of Diabetes, Digestive and Kidney Diseases, Bethesda, MD 20892, USA; Division of Internal Medicine, Krankenhaus Kufstein, Kufstein, Austria; Krankenhaus St. Vinzenz, Zams, Austria; Division of Anesthesia and Intensive Care Medicine, Krankenhaus Meran, Meran, Italy; Department of Anesthesiology, perioperative Medicine and Intensive Care Medicine, Paracelsus Medical University, Salzburg, Austria; Division of Internal Medicine, Krankenhaus St. Johann, St. Johann, Austria; Departments of Oncology & Medicine, Georgetown University, Washington, DC, USA

**Author notes:** equal contribution (Ludwig Knabl, Hye Kyung Lee). Corresponding authors: LK; HKL; PAF; LH.

## Abstract

Fast-spreading variants of severe acute respiratory syndrome coronavirus 2 (SARS-CoV-2) energize the COVID-19 pandemic. The B.1.351 variant carrying the escape mutation E484K in the receptor binding domain is of particular concern due to reduced immunological protection following vaccination. Protection can manifest as early as 10 days following immunization with full protection two weeks following the second dose, but the course is not well-characterized for variants. Here, we investigated the immune transcriptome of six elderly individuals (average age 82 yr.) from an old people’s home, who contracted B.1.351, with four having received the first dose of BNT162b eight to 11 days prior to the onset of COVID-19 symptoms. The patients were hospitalized and received dexamethasone treatment. Immune transcriptomes were established from PBMCs approximately 10 and 35 days after the onset of COVID-19 symptomology. RNA-seq revealed a more intensive immune response in vaccinated patients as compared to unvaccinated ones. Specifically, transcription factors linked to the JAK/STAT pathway, interferon stimulated genes, and genes associated with innate antiviral immunity and COVID-19-SARS-CoV-2 infection were highly enriched in vaccinated patients. This rendered the transcriptomes of the older vaccinated group significantly different than older unvaccinated individuals infected at the same institution and more similar to the immune response of younger unvaccinated individuals (age range 48-62) following B.1.351 infection. All individuals in this study whether vaccinated or not were hospitalized due to B.1.351 infection and one vaccinated patient died illustrating that although an enhanced immune response was documented infection it was insufficient to protect from disease. This highlights the need for maintaining physical distancing and prevention measures throughout the time course of vaccination in older adults.

## Introduction

The SARS-CoV-2 variant B.1.351^1^ has multiple changes in the immunodominant spike protein that facilitates viral cell entry via the angiotensin-converting enzyme-2 (ACE2) receptor. The receptor-binding domain (RBD) mutation E484K provides tighter ACE2 binding and widespread escape from monoclonal antibody neutralization^2-5^, a concern for a limited protection by some vaccines^6^, and recent data from Qatar demonstrate a BNT162b vaccine^7^ effectiveness of 75% with the B.1.351 variant^7^. Vaccine response as well as the immunological response to SARS-CoV-2 infection are time-dependent, with first a rise and then a fall to normal levels of immune-related gene transcription^8^. Differences in sampling times post vaccination and/or infection strongly influence results derived. Limited data is available on the comparative response of the immune transcriptome between infected individuals who are vaccinated versus unvaccinated following the first dose. Since the immune response declines with age, it is most critical to explore this question in the older age group that is most affected by the COVID-19 pandemic. Here, we conducted a study on elderly patients in an old-people’s home that experienced an outbreak of the SARS-CoV-2 B.1.351 variant. A unique aspect of this single setting study is that the infected individuals were all exposed within the same timeframe and vaccinated individuals were all immunized within the same timeframe, controlling for environmental and kinetic variables.

## Results

### Study design

To better understand the extent of protection of elderly individuals from COVID-19 after receiving the first dose of BNT162b vaccine, we investigated a unique cohort of hospitalized patients from an old people’s home that experienced an outbreak of the SARS-CoV-2 variant B.1.351 in January of 2021 with similar kinetics of vaccination, infection and disease development. Six elderly individuals (average age 82 yr.) developed COVID-19 symptoms whole viral genome sequencing confirmed the B.1.351 strain (Supplementary Table 1). Four patients had received the first dose of the BNT162b (Pfizer-BioNTech) vaccine 11 days before the onset of COVID-19 symptoms and one patient eight days prior (Fig. 1a). All patients had underlying health conditions, they were hospitalized and cared for by the same physician and received dexamethasone treatment. Three additional hospitalized B.1.351-positive unvaccinated patients (average age 56) from other communities in Tyrol and South Tyrol (Italy) were included in the broader study (Fig. 1a; Supplementary Table 1). Having this well-controlled cohort of individuals with equivalent ages from the same nursing home, provided the opportunity to explore the impact of the first vaccination on the immune transcriptome in hospitalized COVID-19 patients.

**Fig. 1.**
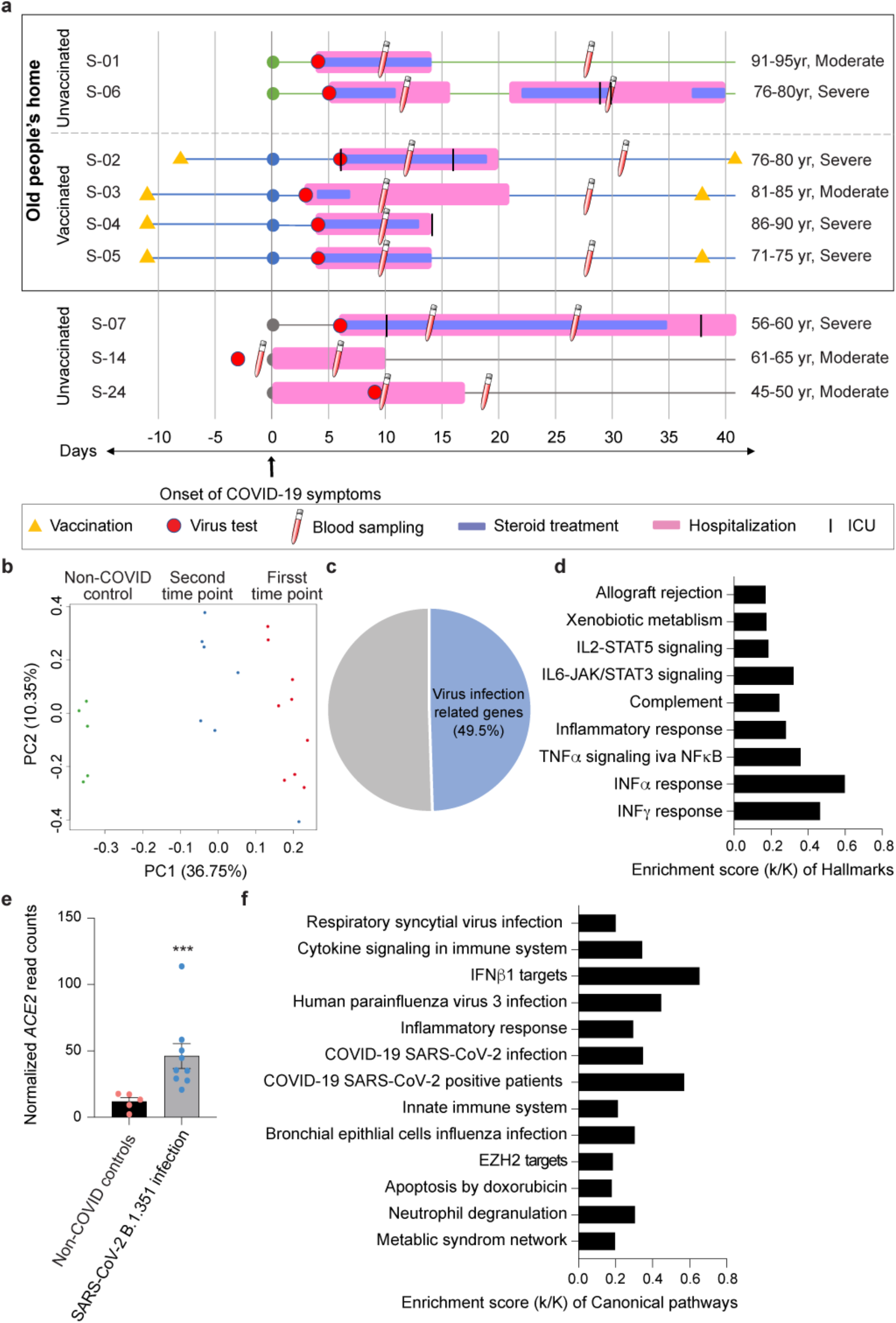
The patient cohort from this study and their immune transcriptomes. **a**. Overview of the patient cohort. Patients S-01 to S-06 live in an old people’s home and developed COVID-19 at the end of January 2021. Patients S-07 to S-09 are from other parts of Tyrol and South Tyrol. Whole viral genome sequencing was used to validate the identity of the B.1.351 variant. Age, COVID-19 severity vaccination status, SARS-CoV-2 test, hospitalization duration, treatment duration and blood sampling time points are shown. For each patient, the day when COVID-19 symptoms were diagnosed was set as 0. S: sample **b**. Principal-component analysis (PCA) of transcriptomes of COVID-19 patients and non-COVID controls^8^, depicting the variation in the global gene expression profiles across the two time points and non-COVID controls. Principal components 1 (PC1) and 2 (PC2), which represent the greatest variation in gene expression, are shown. **c**. Approximately one half of the genes whose expression was induced significantly in COVID-19 patients were linked to pathways activated by virus infection. **d**. Genes expressed at significantly higher levels in the COVID-19 patients were significantly enriched in Hallmark Gene Sets (FDR q value < 0.005). Nine hallmarks that are involved in immune regulation were presented. **e**. Relative normalized expression of ACE2 from COVID-19 patients and healthy controls. \*\*\**p* < 0.0001 by unpaired *t*-test. **f**. Canonical pathway analysis of significantly enriched genes included in clusters for virus infection and innate immune-related biological processes.

### Immune transcriptome

Blood was drawn at ∼10 and 30 days after the development of COVID-19 symptoms, RNA was prepared from buffy coats followed by RNA-seq analyses. The principal component analysis (PCA) showed a separation between RNA-seq samples from non-COVID controls^8^ and the first and second time points from COVID-19 patients on the first principal component (PC1) (Fig. 1b). In total, 3410 genes were significantly differentially expressed between non-COVID controls and the nine COVID-19 patients infected with the B.1.351 variant. The expression of 2026 genes was elevated in COVID-19 patients compared to non-COVID controls (Fig. 1c; Supplementary Table 2). As shown previously^9,10^, the significantly up-regulated genes are enriched in immune response pathways, including IL-JAK/STAT signaling and interferon alpha/gamma responses (Fig. 1d). Expression of the angiotensin-converting enzyme 2 (ACE2) receptor^11,12^ was elevated in the patients (Fig. 1e) suggesting that that the classical or novel dACE2 promoter responded to immune activation^13,14^. Of note, the immune response was linked to viral defense mechanisms (Fig. 1f).

### Elevated immune response in patients infected after the first vaccination

Next, we focused on the immune transcriptome response in six hospitalized elderly patients (average age 82 yr.) from the same old people’s home (Supplementary Table 1), four of which had received the first dose of the BNT162b vaccine 11 days prior to developing COVID-19 symptoms. All six patients developed COVID-19 within two days of each other, were admitted to the same hospital, treated with Fortecortin (Dexamethasone) and were cared for by the same physician. First, we generated the transcriptomes from PBMCs isolated between 9 and 12 days after developing COVID-19 (Fig. 1a; Supplementary Table 1). The PCA plot shows a separation between the vaccinated and unvaccinated groups on PC1 (Fig. 2a), which is further strengthened by hierarchical clustering of the 181 significantly differentially expressed genes between the six patients (Fig. 2b; Supplementary Table 3). Of the 85 genes significantly upregulated in vaccinated patients, 72% (61 genes) are enriched in anti-viral immune responses and influenza vaccination^15^, such as IFN α/γ responses, complement and IL6-JAK/STAT3 signaling pathways (Fig. 2c-d; Supplementary Table 3). Interferon stimulated genes (ISGs), and genes associated with innate antiviral immunity and COVID-19-SARS-CoV-2 infection are highly enriched in vaccinated patients (red dots) (Fig. 2e; Supplementary Fig. 1; Supplementary Table 3). Specifically, the highly ranked ETV7, STAT1 and STAT2 (Fig. 2f) are key transcription factors executing interferon responses and are top predictors for ACE2 expression in human airway epithelium^14,16-20^. Notably, expression of *IFITM3*, a gene with a polymorphism that has been linked to the severity of COVID-19^21^, is highly induced in vaccinated patients (Supplementary Fig. 1a). While three out of the four vaccinated patients had a highly activated immune transcriptome, patient S-02 had a blunted immune response, more similar to that seen in the non-vaccinated patients (Supplementary Fig. 1b). One explanation is the shorter time period between vaccination and the onset of symptoms.

**Figure 2.**
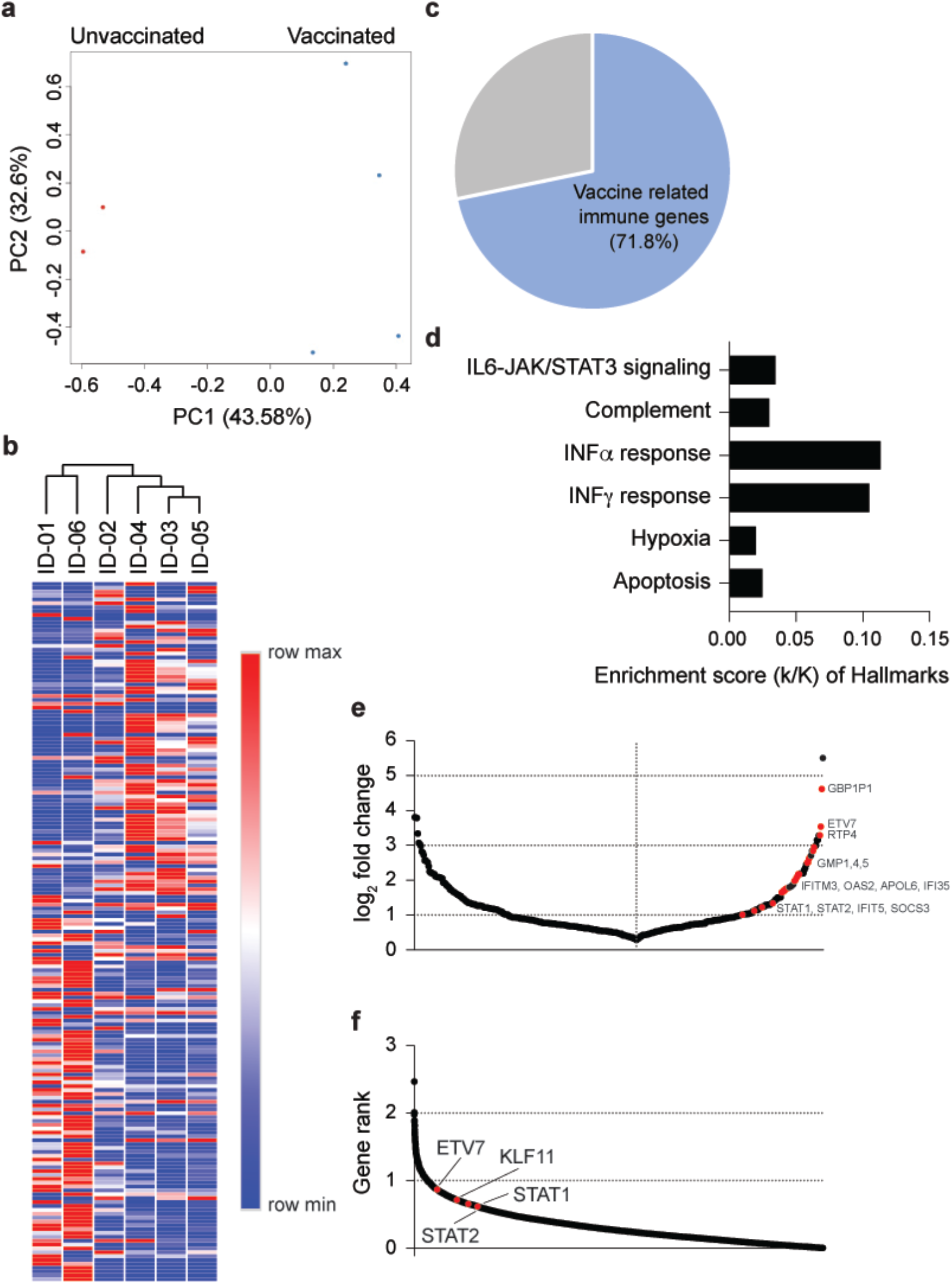
Differences of immune response between non-vaccinated and vaccinated COVID-19 patients. **a**. PCA plot of COVID-19 samples from the six elderly patients from same nursing home on the basis of the expression of all genes. Samples are color-coded by vaccination. **b**. Heatmap shows the expression levels of significantly differentially expressed genes of the samples. Each sample is hierarchically clustered by using their adjacency scores as distance. **c**. 71.8% of the genes induced significantly in the four vaccinated patients infected by the B.1.351 variant are associated with immune responses activated by vaccines and virus infection. **d**. Genes expressed at significantly higher levels in the vaccinated patients were significantly enriched in Hallmark Gene Sets (FDR q value < 0.005). Of six, four hallmarks are related to immune regulation. **e**. Dot plot of DEGs comparing non-vaccinated and vaccinated patients. DEGs (padj < 0.05) with a log_2_(fold change) are indicated and significant genes are in red. **f**. Transcription factors ranked according to association strength with ACE2 expression (for details see Methods). Top 10 hits are highlighted.

### Temporal progression of the immune response

From our cohort of the four vaccinated patients, one died of COVID-19 and three were discharged from the hospital after a single stay of 10-14 days (Fig. 1a). From the two non-vaccinated patients, one was discharged after a single ten day stay and one was re-admitted. To dig deeper into the temporal immune response after discharge from the hospital we analyzed the immune transcriptome of the three vaccinated patients approximately three weeks after the first transcriptome analysis (Figs. 1a and 3; Supplementary Table 4). Gene enrichment analyses of differentially expressed transcripts illustrate a greatly diminished immune response, including INFα/γ signaling, in the recovered patients (Fig. 3a; Supplementary Table 4). Expression of specific ISGs (e.g., IFI35, IFIT5, IFITM3, STAT1, STAT2, OAS2) was sharply reduced (Fig. 3b), approaching levels seen in non-COVID samples.

**Figure 3.**
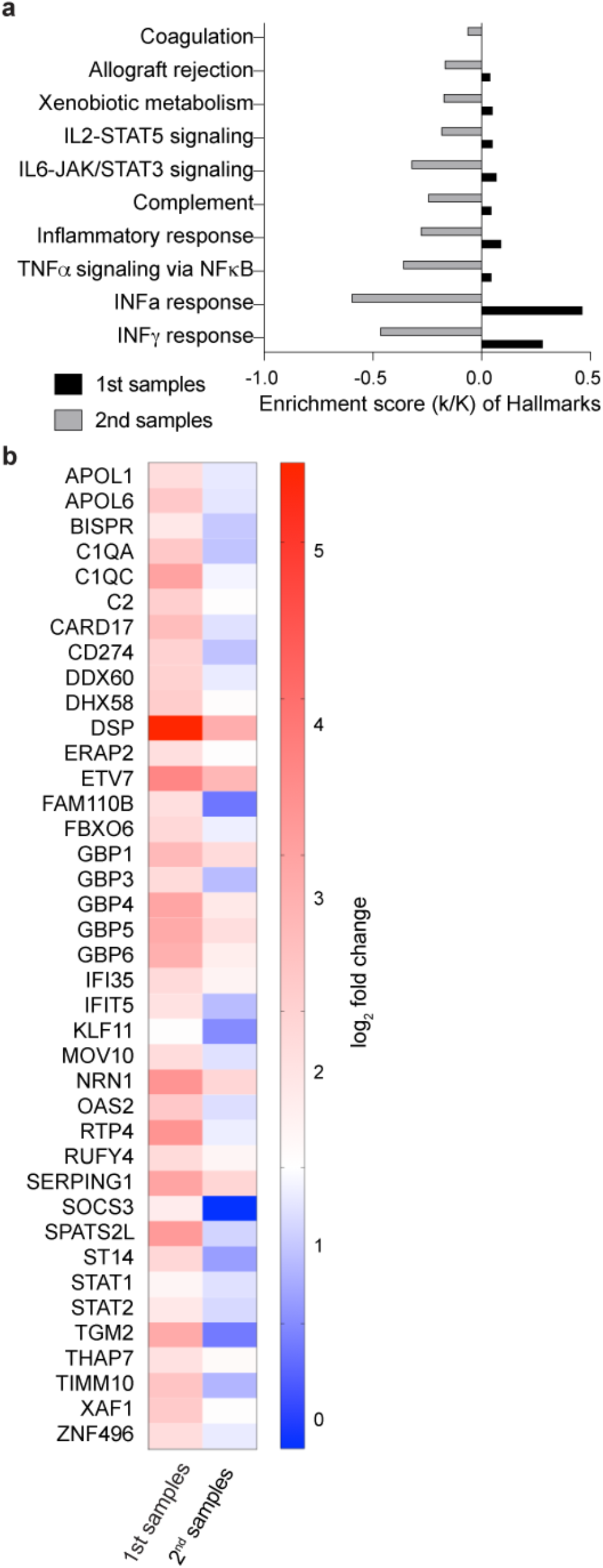
Longitudinal Analysis of the COVID-19 patients in the cohort. **a**. Expression of immune pathway genes induced at the early stage of symptom onset was mitigated after approximately 30 days of symptom onset. **b**. Heatmaps showing relative expression level for a subset of significant genes that are induced in the 1^st^ blood samples and reduced in 2^nd^ samples. Genes included have a log_2_(fold change) of more than 2 and a padj value of less than 0.05.

### Immune responses at different ages

Having analyzed the immune transcriptome from the elderly hospitalized population (average age 82 yr.) we addressed potential age differences and included RNA-seq transcriptomes from three hospitalized patients all below 62 years of age (average age 56 yr.) (Fig. 1a). The two groups were separated in the PCA plot (Fig. 4a; Supplementary Table 5). The immune response in the three younger patients exceeded that of the elderly population and of the 417 significant differentially activated genes, most are related to immune responses and virus infection pathways (Fig. 4b-c; Supplementary Table 5). These results support the concept that age might be a defining factor in the immune response in COVID-19 patients.

**Figure 4.**
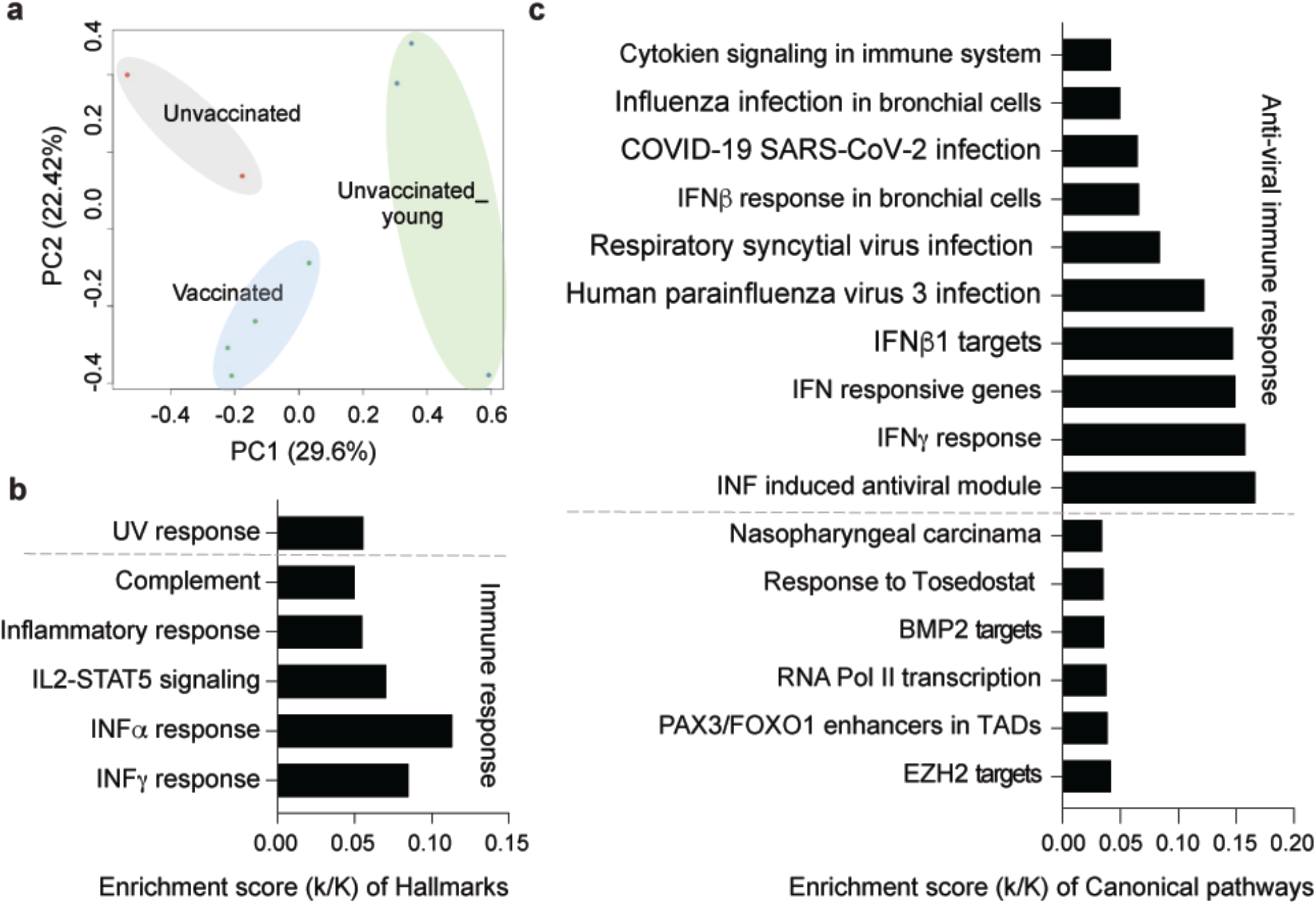
Comparative analysis of the immune response at different age groups. **a**. Sparse PCA depicting global transcriptional profiles of all nine samples at the first time point (see Fig. 1a). **b-c**. Box plots showing hallmark and canonical pathway analysis enriched in DEGs. Most hallmarks of transcripts were included in innate immune-related biological processes. More than half of significant genes are enriched anti-viral immune responses.

## Discussion

COVID-19 is a rapidly evolving field in which real-world data, especially those concerning the now prevalent variants of concern (VOC), is critical for everyday management of the COVID-19 disease by public health officials, government agencies and institutions. There have been no studies of whole transcriptome response following first vaccine in infected versus uninfected individuals.

It is known that the first vaccine dose is not fully protective^22^ but data available indicate that as early as 12 days following first vaccination there may be a response against the dominant variants B.1.1.7 and B.1.351^7,23^. While that may be reassuring in large population studies, here we show that for elderly patients, reflective of groups with higher inherent risk of morbidity and mortality from SARS-CoV-2 infection, the first vaccine provided minimal clinically significant protection if any. We did not observe a difference in disease severity between the vaccinated and unvaccinated groups and one of the vaccinated individuals died. Our research underscores the difference between public health studies, which have relevance for population statistics, and individual patient management where our data underscores the need for maintenance of strict social distancing and infection control measures throughout the vaccination process.

Our work documented a stronger immune response in patients that had received their first dose of BNT162b prior to infection but no apparent clinical benefit. Breakthrough SARS-CoV-2 infection following full vaccination also occurs but it may present as asymptomatic or milder disease^24,25^. Future research directed at how vaccination influences immune response following infection may help elucidate molecular underpinnings of SARS-CoV-2 disease presentation and contribute to an understanding how best to manage timing of third boosters and variant targeting.

Our study illustrates that whole transcriptome study from buffy coat is feasible and yields actionable data when performed in well-controlled clinical settings. It demonstrates a technique that is feasible to expand and reproduce across settings. While our study was limited to infection ∼11 days following first vaccination, extensions could compare the immune transcriptome after the second vaccination and investigate individuals who may have waning antibody response ten or more months following either infection and/or full immunization. As a caveat, it can be challenging to reliably compare RNA-seq data generated on different platforms and the disease severity at the time of the analysis greatly impacts the transcriptome^10^.

While there was a measurable immune transcriptome response to first vaccination, it was insufficient to prevent significant clinical disease and protective measures against spread should be fully employed in individuals who have received only one dose. Moreover, SARS-CoV-2 variants of concern (VOC) such as B.1.351 studied here demonstrate resistance to neutralization by antibodies generated from some current vaccines^11^. Further research in vaccinology will continue to address mechanisms to improve the protective immunological response to SARS-CoV-2 and VOC through boosters and vaccines engineered specifically for VOC. Serial immune transcriptome studies should be included in addition to antibody tests for a fuller understanding of the spectrum of immune response in real-world situations as more immunized individuals encounter SARS-CoV-2 VOC.

### Limitations

The findings in this report are subject to at least three limitations. We have investigated the transcriptome from patients infected with B.1.351 but not with other variants. Our study focused on elderly (average age 82 yr.) vaccinated and unvaccinated patients with limited comparison to younger individuals (48-62 yr). We have investigated the effect of the BNT162b vaccine but not of any other vaccine. The study group was small; however, the parallel timing of vaccination, infection and treatment in the elderly group enabled the study to be well controlled for environmental factors.

## Methods

### SARS-CoV-2 virus sequencing

RNA was extracted from patient’s blood using a Maxwell RSC simply RNA Blood purification kit according to the manufacturer’s instructions (Promega, USA). Library preparation and sequencing was performed as described^26^. In short, cDNA was obtained by using reverse transcriptase with random priming. Following cDNA synthesis, primers based on sequences from the ARTICnetwork were used to generate 400 bp amplicons in two different PCR pools. After merging of pools and amplification, libraries were constructed using QIASeq FX DNA Library UDI Kit following the manufacturer’s instructions (Qiagen GmbH, North Rhine-Westphalia, Germany).

Sequencing was done with Illumina NextSeq® 500/550 using 149-bp paired-end reads with 10-bp indices (Illumina, California, USA). Obtained viral sequences were assembled using CLC Genomics Workbench v20.0.3 (Qiagen GmbH, North Rhine-Westphalia, Germany). SARS-CoV-2 isolate Wuhan-Hu-1 served as the reference genome (Accession NC_045512.2). SARS-CoV-2 variants were identified by uploading FASTA-files on freely accessible databases (http://cov-lineages.org/).

### Extraction of the buffy coat and purification of RNA

Whole blood was collected, and total RNA was extracted from the buffy coat and purified using the Maxwell RSC simply RNA Blood Kit (Promega) according to the manufacturer’s instructions. The concentration and quality of RNA were assessed by an Agilent Bioanalyzer 2100 (Agilent Technologies, CA).

### mRNA sequencing (mRNA-seq) and data analysis

The Poly-A containing mRNA was purified by poly-T oligo hybridization from 1 μg of total RNA and cDNA was synthesized using SuperScript III (Invitrogen, MA). Libraries for sequencing were prepared according to the manufacturer’s instructions with TruSeq Stranded mRNA Library Prep Kit (Illumina, CA, RS-20020595) and paired-end sequencing was done with a NovaSeq 6000 instrument (Illumina).

The raw data were subjected to QC analyses using the FastQC tool (version 0.11.9) (https://www.bioinformatics.babraham.ac.uk/projects/fastqc/). mRNA-seq read quality control was done using Trimmomatic^27^ (version 0.36) and STAR RNA-seq^28^ (version STAR 2.5.4a) using 150 bp paired-end mode was used to align the reads (hg19). HTSeq^29^ (version 0.9.1) was to retrieve the raw counts and subsequently, R (https://www.R-project.org/), Bioconductor^30^ and DESeq2^31^ were used. Additionally, the RUVSeq^32^ package was applied to remove confounding factors. The data were pre-filtered keeping only genes with at least ten reads in total. The visualization was done using dplyr (https://CRAN.R-project.org/package=dplyr) and ggplot2^33^. Genes were categorized as significant differentially expressed with an adjusted p-value (pAdj) below 0.05 and a fold change > 2 for up-regulated genes and a fold change of < −2 for down-regulated ones and then conducted gene enrichment analysis (https://www.gsea-msigdb.org/gsea/msigdb).

### Statistical analysis

Data were presented as the means ± s.e.m. (standard error of the mean) of all experiments with *n* = number of biological replicates. For comparison of RNA expression levels between two groups, data were presented as standard deviation in each group and were evaluated with a two-way ANOVA followed by Tukey’s multiple comparisons test or a two-tailed unpaired t-test with Welch’s correction using GraphPad PRISM (version 9.0). A value of \**P* < 0.05, \*\**P* < 0.001, \*\*\**P* < 0.0001, \*\*\*\**P* < 0.00001 was considered statistically significant.

## Data Availability

The RNA-seq data of patients will be uploaded in GEO before publishing the manuscript.

## Ethics approval

This study was approved by the Institutional Review Board (IRB) of the Office of Research Oversight / Regulatory Affairs, Medical University of Innsbruck, Austria (EC numbers: 1064/2021).

## Data availability

The RNA-seq data of patients will be uploaded in GEO before publishing the manuscript.

## Acknowledgments

Our gratitude goes to the patients who contributed to this study to advance our understanding of COVID-19. This work was supported by the Intramural Research Programs (IRPs) of National Institute of Diabetes and Digestive and Kidney Diseases (NIDDK) and utilized the computational resources of the NIH HPC Biowulf cluster (http://hpc.nih.gov). RNA-sequencing was conducted in the NIH Intramural Sequencing Center, NISC (https://www.nisc.nih.gov/contact.htm).

## Author contribution

LK: recruited patients, collected material, analyzed data; HKL: analyzed data, wrote manuscript; MW: collected and prepared material; AM, AZ, LK Sr, SR, MB, JS and NK: recruited and diagnosed patients; PAF: analyzed data, wrote manuscript; LH: analyzed data, wrote manuscript. All authors read and approved the manuscript

## Figure legends

**Supplementary Figure 1.**
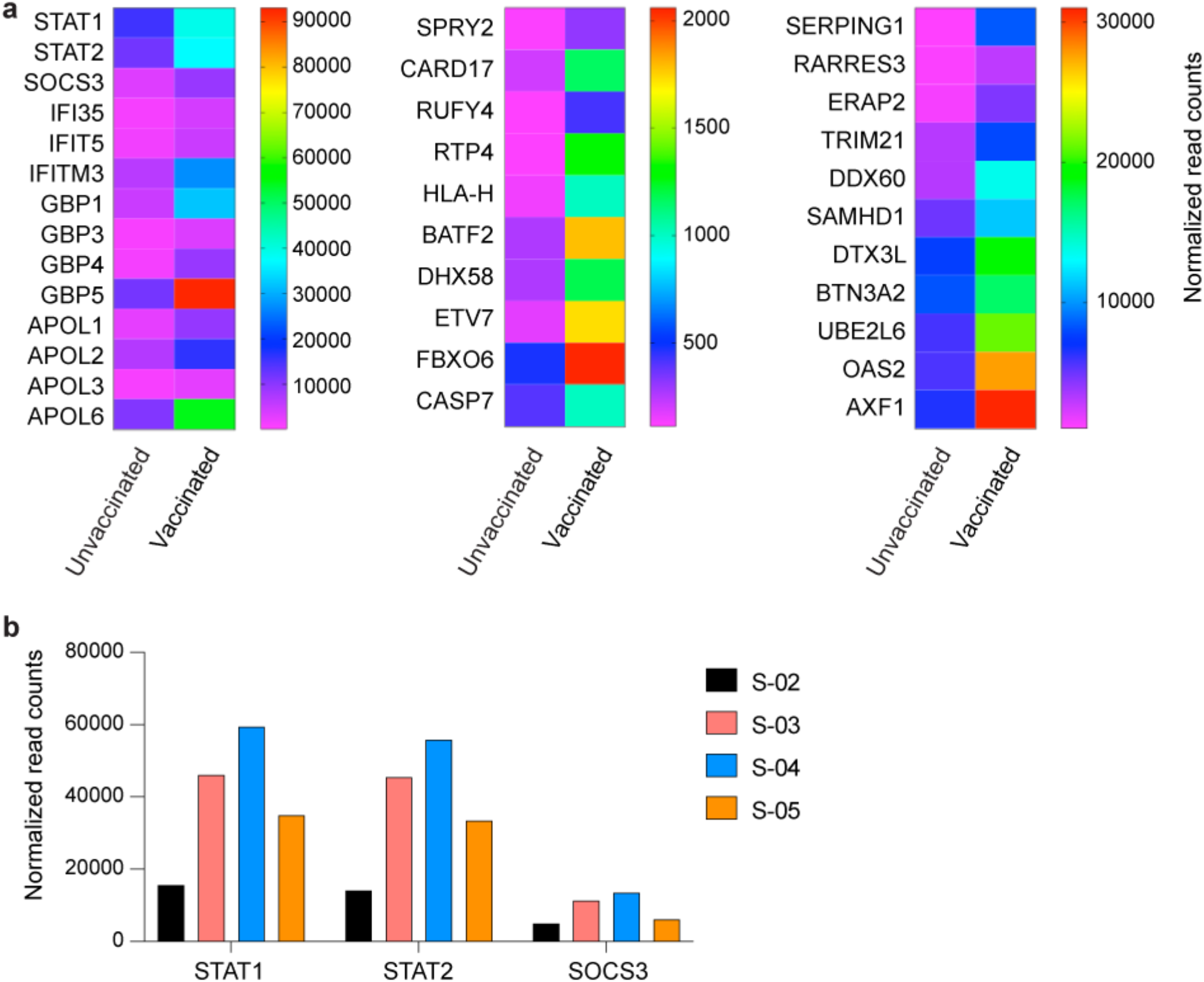
Upregulation of Interferon stimulated genes (ISGs), innate antiviral immunity and COVID-19-SARS-CoV-2 infection genes in vaccinated COVID-19 patients. **a**. Heatmap showing the significant enriched genes related to immune responses and SARS-CoV-2 virus infection. **b**. mRNA levels of JAK/STAT signaling components, STAT1, STAT2 and SOCS3, measured by RNA-seq were presented by bar graphs. S: sample

